# Multiple mechanisms of strain competition are needed to explain pneumococcal serotype-specific antibiotic resistance patterns

**DOI:** 10.1101/2025.05.16.25327750

**Authors:** Hannah C. Lepper, Emma Pujol-Hodge, Jada Hackman, Kevin van Zandvoort, Kate Mellor-Wright, Oliver Lorenz, Stephanie Lo, Duc Anh Dang, Hung Thai Do, Gerry Tonkin-Hill, Stephen D. Bentley, Lay Myint Yoshida, Stefan Flasche, Nicholas G. Davies, Katherine E. Atkins

## Abstract

2

**Background:** Disease caused by antibiotic resistant pneumococcal strains is an important public health concern. The impact of public health interventions depend on the complex ecology governing serotype and antibiotic resistant strain dynamics, for which we currently lack mechanistic explanations. Ee evaluated whether multi-serotype transmission models of antibiotic resistant pneumococcal strains can recapitulate empirical resistance data, thus providing a plausible explanation for how antibiotic-sensitive and resistant strains coexist at intermediate frequencies, both between and within serotypes.

**Methods:** We extracted a global dataset of pneumococcal carriage prevalence in healthy, unvaccinated children by serotype and penicillin susceptibility from a previously-published systematic review. Next, we developed a suite of multi-serotype individual-based models that have previously been shown to plausibly explain coexistence in single serotype models, incorporating transmission and clearance rate differences between serotypes and serotype-specific immunity. Finally, we calibrated the suite of models to the relative serotype prevalence and overall resistance frequency drawn from the extracted dataset.

**Results:** Overall resistance frequency varied considerably between studies, but we found a positive association between frequency of a serotype and resistance frequency within that serotype, and a high degree of coexistence of sensitive and resistant strains within serotypes. Each individual-based model predicted different serotype-specific resistance frequencies, but combining models was necessary to capture all features of the empirical data.

**Conclusions:** Serotype-specific resistance patterns may give important clues to understand fundamental antibiotic resistance epidemiology. We show that combining multiple independently plausible mechanisms can capture resistant pneumococcal carriage, but additional data are needed to determine the strength of these individual mechanisms within these combinations.

## 3 Introduction

Antibiotic-resistant bacterial infections are estimated to kill around 1.14 million people annually (Murray et al., 2022; Naghavi et al., 2024). Designing interventions that effectively reduce drug resistant infections is therefore a global priority (Vekemans et al., 2021). Predicting the impact of an intervention aiming to reduce resistance requires first understanding which factors drive the frequency with which bacteria are resistant to antibiotics in the absence of an intervention. However, finding such a mechanistic explanation has been a fundamental challenge (Lipsitch et al., 2009; Davies et al., 2021). Moreover, for many bacterial species that cause substantial antibiotic resistant deaths, such as *Streptococcus pneumoniae* and *Escherichia coli*, explaining the stable intermediate frequency of resistance becomes harder, because the species have multiple serotypes which themselves differ in their level of antibiotic resistance (Andrejko et al., 2022; Weerdenburg et al., 2023). As we currently lack a mechanistic understanding of how antibiotic resistance is stably maintained across bacterial serotypes, we have limited ability to predict the impact of interventions designed to reduce disease and deaths from antibiotic resistance. In *S. pneumoniae*, which accounts for an estimated 10% of deaths due to antibiotic resistant infections (Murray et al., 2022; Naghavi et al., 2024), resistance is more frequent in the most prevalent serotypes, which cause the most invasive disease (Lo et al., 2019; Andrejko et al., 2022; Song et al., 2012). There are over 100 different serotypes (Ganaie et al., 2020), each of which has a different polysaccharide capsule that elicits serotype-specific immune responses (Hausdorff et al., 2005). Serotype also differ in traits such as carriage duration which could impact selection for resistance (Lehtinen et al., 2017; Lees et al., 2017).

Some serotypes are more likely to cause invasive disease than others, and pneumococcal conjugate vaccines (PCVs) protect against transmission and disease from a subset of virulent serotypes (Gallagher et al., 2025; Lewnard and Hanage, 2019). PCV has been proposed as a key intervention for reducing antibiotic resistance, and overall resistance frequency in pneumococcal carriage and infection has been observed to reduce after vaccine introduction (Andrejko et al., 2021). However, resistance frequency to some antibiotics has also increased in non-PCV serotypes (Lo et al., 2019; Andrejko et al., 2021), suggesting complex dynamics may be at play. To capture pneumococcal resistance epidemiology and predict the impact of interventions, it is important to be able to explain how resistance is selected for at the serotype level.

Several microbial ecological mechanisms could explain the co-occurrence of sensitive and resistant strains of the same bacterial species in host populations (Lehtinen et al., 2017; Davies et al., 2019; Blanquart et al., 2018; Krieger et al., 2020; Davies et al., 2021; Colijn et al., 2010). However, we still do not have a coherent understanding of how these mechanisms interact with serotype dynamics to generate empirical patterns of pneumococcal resistance. To address this, we built a dataset of global serotype-specific resistance patterns in pneumococcal carriage in healthy children before PCV introduction, and developed a suite of multi-serotype individual-based mathematical models for each mechanism. By comparing predictions of each model individually and in combination to our dataset, we assessed the role of each mechanism in shaping serotype-level selection for resistance.

## 4 Results

### 4.1 Serotype-specific resistance patterns in pneumococcal carriage studies

To build a dataset of global serotype-specific resistance patterns, we systematically identified studies reporting serotype-specific prevalence and penicillin susceptibility among studies included in an existing systematic review Andrejko et al. (2021). Of the 558 studies screened, we found 12 studies that met all inclusion criteria (S. Fig 3.), covering 9,977 children from 12 countries between 1995 and 2019 (Supplementary Materials). The average pneumococcal carriage prevalence across all samples was 35.5%, with study prevalences ranging from 14.1% in Singapore in 2008 to 78.4% in Norway in 2006, which the authors identify as an unusually high prevalence for this setting Vestrheim et al. (2008). In total, 50 serotypes were identified, with 41.4% of all isolates resistant or non-susceptible to penicillin, with individual study frequencies ranging from 3.7% in Sweden in 2005 to 93.5% in Jordan in 2019.

We applied mixed-effects binomial regression models to describe serotype-specific resistance patterns in the data. Under the best-fitting model, we found a significant (p=0.03) positive association between serotype frequency rank and resistance frequency (7% increase in the odds of non-susceptibility per unit increase in rank) (S.Table 4). However, over half of the variance in the observed data (52%) was explained by differences between studies, indicating that other unobserved factors associated with region affect resistance frequency of a serotype. Serotype identity itself only explained 4% of variation in the data while serotype rank as a fixed effect explained 2% (S.Table 4).

Coexistence of resistant and sensitive strains of the same serotype was observed in all studies. In the study with the most samples, Espinosa-de los Monteros et al. (2007), 94.1% of serotypes with more than five or more isolates had an intermediate resistance frequency (between 5 and 95%). However, in many studies there were low number of isolates or a low resistance frequency, making serotype-specific resistance frequency within that study uncertain (Fig 1).

**Figure 1:**
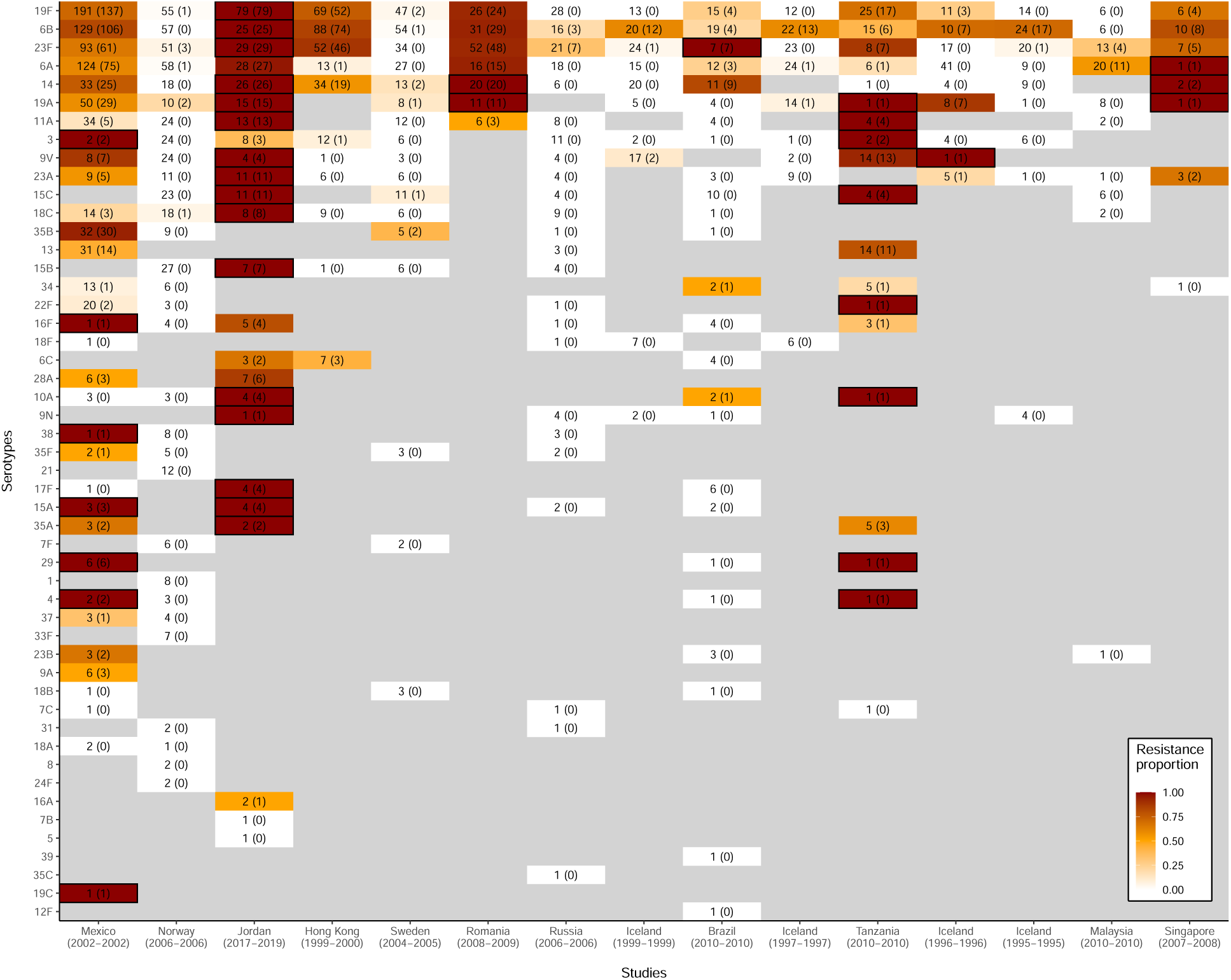
Resistance frequency varied by study, serotype, and serotype rank in the empirical data. Total *S. pneumoniae* isolates and non-susceptible isolates (in brackets) for each study and serotype. The colour of the tiles indicates the relative non-susceptible frequency for the serotype, with black-outlined boxes indicating cases of 100% non-susceptible frequency. Studies on the x-axis are in decreasing order of total number of isolates analysed, while serotypes on the y-axis are in decreasing order of total prevalence.

**Figure 2:**
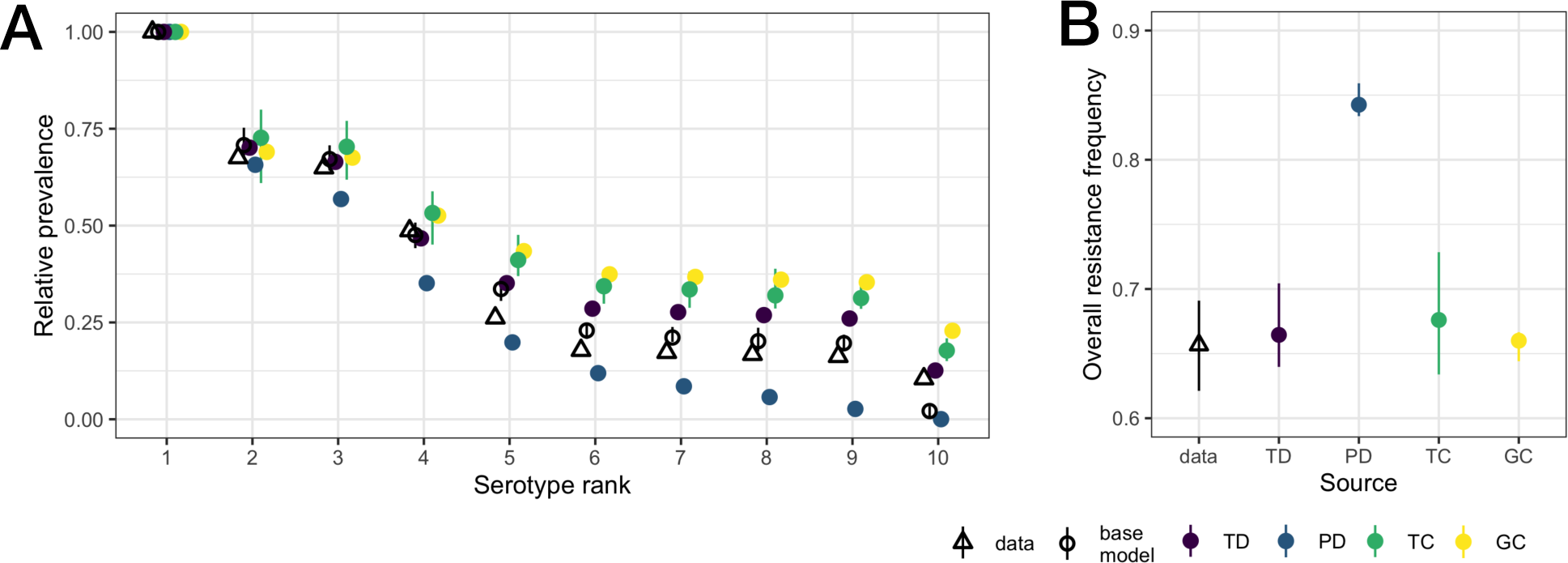
model fitting results.

### 4.2 Serotype-specific resistance patterns predicted by mathematical models

To evaluate our ability to recapitulate these empirical data, we developed a suite of four individual-based transmission models for each of the coexistence mechanisms that can plausibly reproduce non-serotype specific resistance frequency identified in Davies et al. (2021) (Table 1). We then used data on relative serotype prevalence and overall resistance frequency from the extracted dataset to calibrate the models.

**Table 1:**
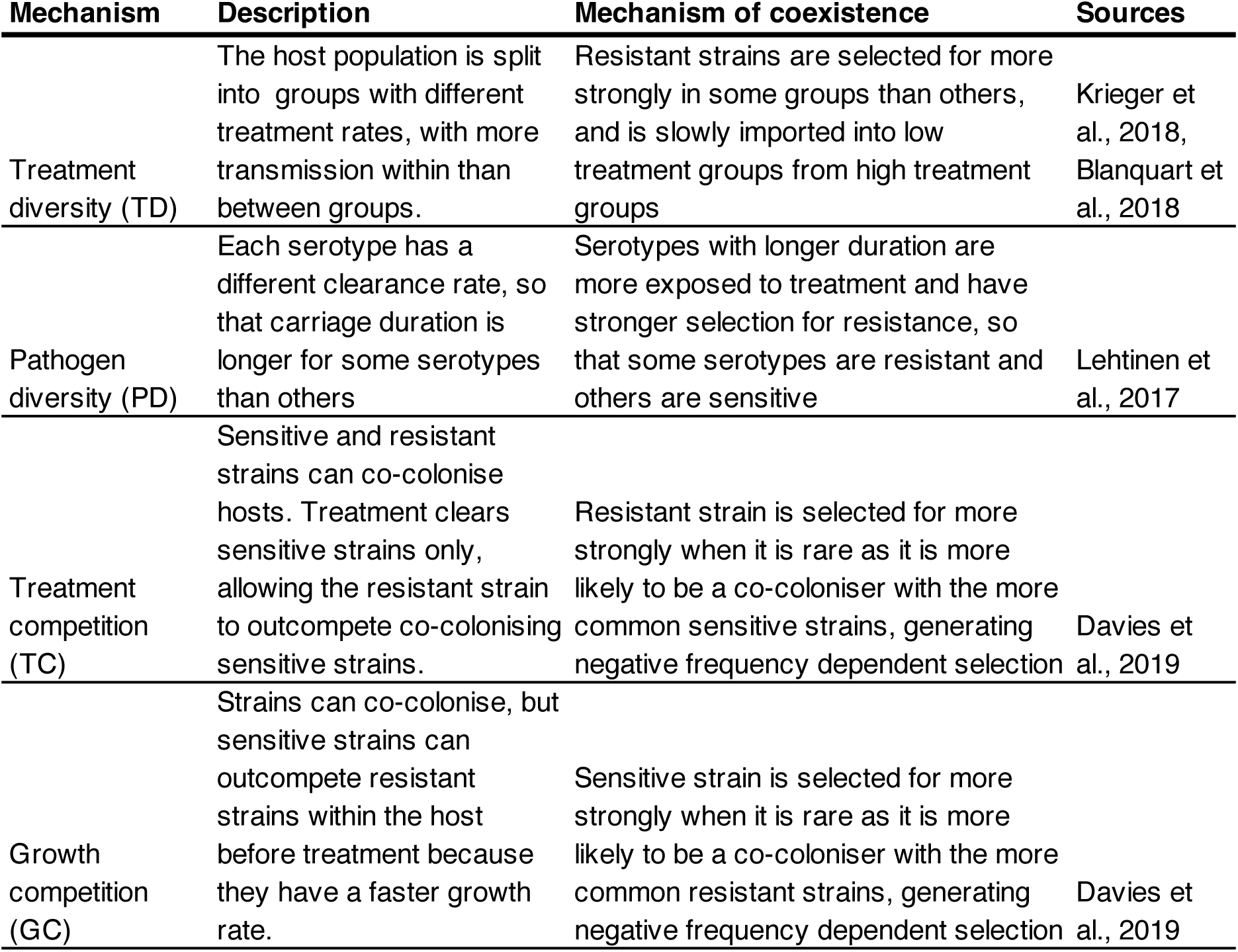
Description of each modelled mechanism. Drawn from (Davies et al., 2019).

| **Mechanism** | **Description** | **Mechanism of coexistence** | **Sources** |
| --- | --- | --- | --- |
| Treatment diversity (TD) | The host population is split into groups with different treatment rates, with more transmission within than between groups. | Resistant strains are selected for more strongly in some groups than others, and is slowly imported into low treatment groups from high treatment groups | Krieger et al., 2018, Blanquart et al., 2018 |
| Pathogen diversity (PD) | Each serotype has a different clearance rate, so that carriage duration is longer for some serotypes than others | Serotypes with longer duration are more exposed to treatment and have stronger selection for resistance, so that some serotypes are resistant and others are sensitive | Lehtinen et al., 2017 |
| Treatment competition (TC) | Sensitive and resistant strains can co-colonise hosts. Treatment clears sensitive strains only, allowing the resistant strain to outcompete co-colonising sensitive strains. | Resistant strain is selected for more strongly when it is rare as it is more likely to be a co-coloniser with the more common sensitive strains, generating negative frequency dependent selection | Davies et al., 2019 |
| Growth competition (GC) | Strains can co-colonise, but sensitive strains can outcompete resistant strains within the host before treatment because they have a faster growth rate. | Sensitive strain is selected for more strongly when it is rare as it is more likely to be a co-coloniser with the more common resistant strains, generating negative frequency dependent selection | Davies et al., 2019 |

To capture a single epidemiological setting, we selected the largest dataset extracted from the systematic review, Espinosa-de los Monteros et al. (2007), which reported 2777 samples from 12 states in Mexico, extracting the frequencies of the 10 most prevalent serotypes and the overall penicillin resistance rate (65.7%) (2B).

To test the performance of these calibrated models, we then compared each model output with three key qualitative observations generated from the entire systematic review dataset: 1) overall population-level coexistence of sensitive and resistant strains (of any serotype); 2) a positive association between serotype frequency and resistance frequency; 3) and high within-serotype coexistence, i.e. a high proportion of serotypes maintaining an intermediate resistance frequency.

Each individual model generated a range of stable intermediate frequencies of resistance with increasing treatment rate when incorporating multiple serotypes, consistent with results from the respective single-serotype models (S. Fig 4). However, each model generated a different serotype-specific resistance pattern, and no single model could simultaneously capture both the positive serotype frequency-resistance frequency association and intermediate proportions of resistance frequency within serotypes. Under the treatment diversity model, every serotype had the same resistance frequency, with increasing variance with decreasing serotype rank (Fig 3A, Table 2). Consistent with previous studies (Lehtinen et al., 2017), the pathogen diversity model predicted that serotypes above the carriage duration threshold are all resistant, and those below are all sensitive (Fig 3A, Table 2). Both the treatment and growth competition models predicted that serotypes tend to have close to either 0% or 100% resistance frequency, with intermediate resistance frequency within a serotype being rare (Fig 3A, Table 2). Whereas resistance frequency was positively associated with serotype prevalence in the treatment competition model, the growth competition model resulted in the opposite pattern (Fig 3A, Table 2). We investigated why there was a lack of intermediate resistance frequencies within serotypes in the within-host models with a simplified, 2-serotype version of the model and invasion experiments, which indicated that immune competition can prevent both strains of the same serotype from coexisting (described in Supplementary Materials).

**Figure 3:**
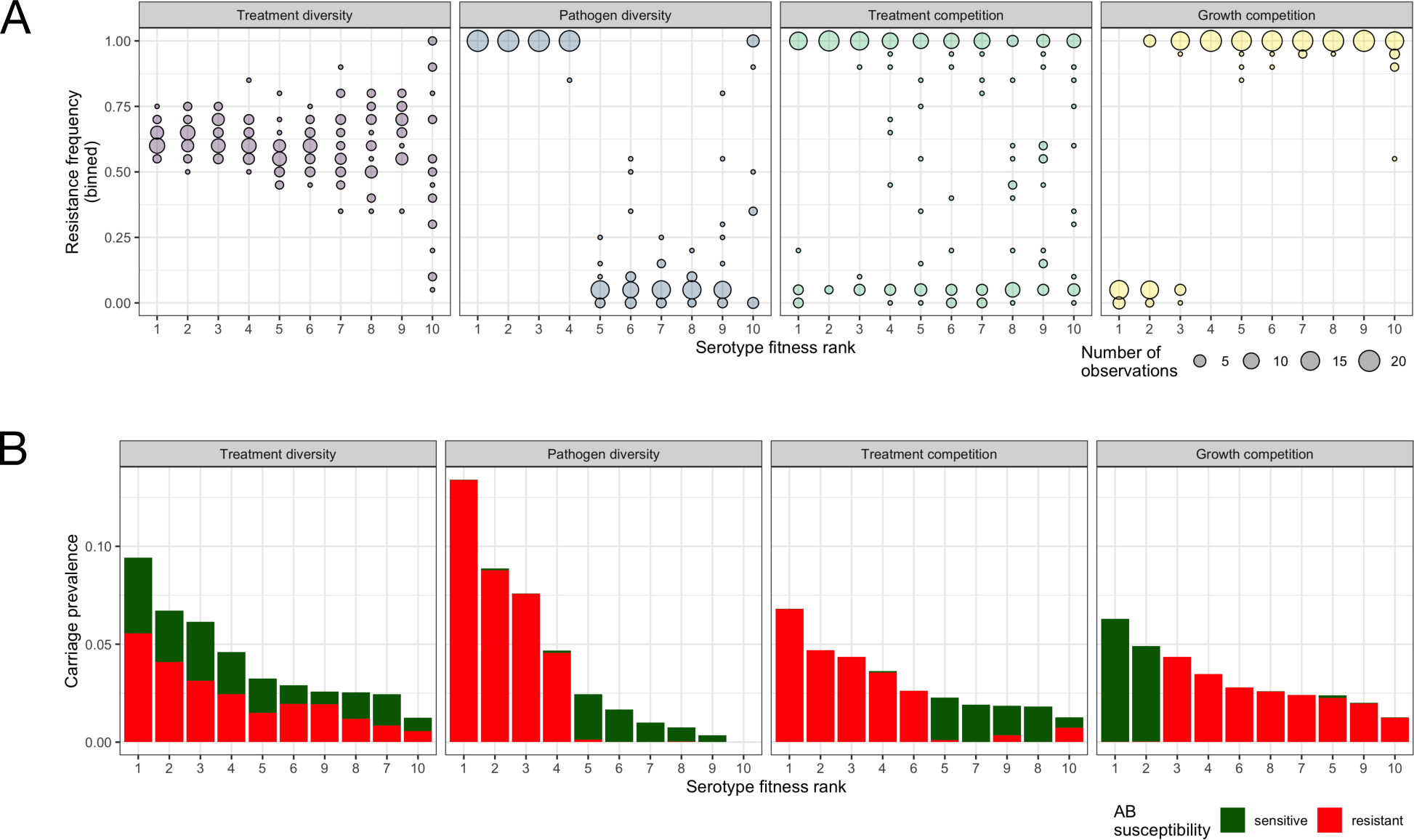
Each mechanism predicts a different serotype-specific resistance pattern. A) Variation in resistance frequency of each hypothetical serotype from 20 model simulations. Resistance frequency (y axis) is binned, with the size of the circle indicating the number of repeated model simulations that fell within that bin. B) Example carriage prevalence of the 10 most prevalent serotypes for a single simulation of each model. Red indicates the proportion of samples of a given serotype that were non-susceptible to penicillin and green represents the proportion that were sensitive.

**Table 2:** Only combined models can capture all qualitative characteristics in the empirical data. Comparison of serotype-specific resistance pattern characteristics in the data and the model. Spearman’s Rho gives the correlation between serotype fitness rank and resistance frequency rank within each repeated model simulation, with 95% confidence intervals. Within-serotype coexistence proportion gives the average proportion of serotypes that had intermediate resistance frequencies (between 5 and 95%) across model simulations, with 95% quantiles.

| **Model** | **Captures positive relationship between serotype prevalence and resistance frequency?** | **Captures high levels of within-serotype coexistence?** |
| --- | --- | --- |
| *Individual models:* |  |  |
| Treatment diversity (TD) | No (0.03, -0.10 to 0.17) | Yes (0.98, 0.96 to 1.00) |
| Pathogen diversity (PD) | Yes (0.62, 0.52 to 0.70) | No (0.20, 0.16 to 0.25) |
| Treatment competition (TC) | Yes (0.22, 0.09 to 0.36) | No (0.26, 0.20 to 0.31) |
| Growth competition (GC) | No (-0.38, -0.58 to -0.25) | No (0.02, 0.00 to 0.03) |
| *Combined models:* |  |  |
| PDxTD | Yes (0.86, 0.82 to 0.89) | Yes (0.52, 0.47 to 0.56) |
| PDxTC | Yes (0.66, 0.58 to 0.73) | No (0.12, 0.12 to 0.24) |
| PDxGC | No (-0.04, -0.18 to 0.10) | No (0.20, 0.15 to 0.24) |
| TDxTC | Yes (0.64, 0.56 to 0.72) | Yes (0.99, 0.96 to 1.00) |
| TDxGC | No (-0.80, -0.85 to -0.74) | Yes (0.69, 0.65 to 0.74) |
| PDxTDxTC | Yes (0.89, 0.86 to 0.92) | Yes (0.50, 0.46 to 0.54) |
| PDxTDxTCxGC | Yes (0.87, 0.83 to 0.90) | Yes (0.49, 0.43 to 0.54) |

We then created two-, three- and four-way combinations of each mechanism to test whether observed serotype-specific patterns were obtainable when multiple drivers of resistant-sensitive strain competition were operating simultaneously (Fig 4). Adding the treatment diversity mechanism to any other model increased within-serotype coexistence, without affecting the predicted relationship between serotype and resistance frequency (Table 2, Fig 4A). The treatment competition or the pathogen diversity models could still capture positive serotype frequency-resistance frequency relationships when combined either together or with the treatment diversity model (Table 2, Fig 4A). However the growth competition model performed poorly in all 2-way combinations, predicting a negative relationship between serotype and resistance frequencies (Fig 4A). Combining the treatment diversity, treatment competition, and pathogen diversity mechanisms also agreed with observed patterns (Table2, Fig 4B). We did not combine the treatment and growth competition models in a 2-way combination because this would have resulted in two different resistance costs in the same model, and would have required additional fitting to be meaningful.

**Figure 4:**
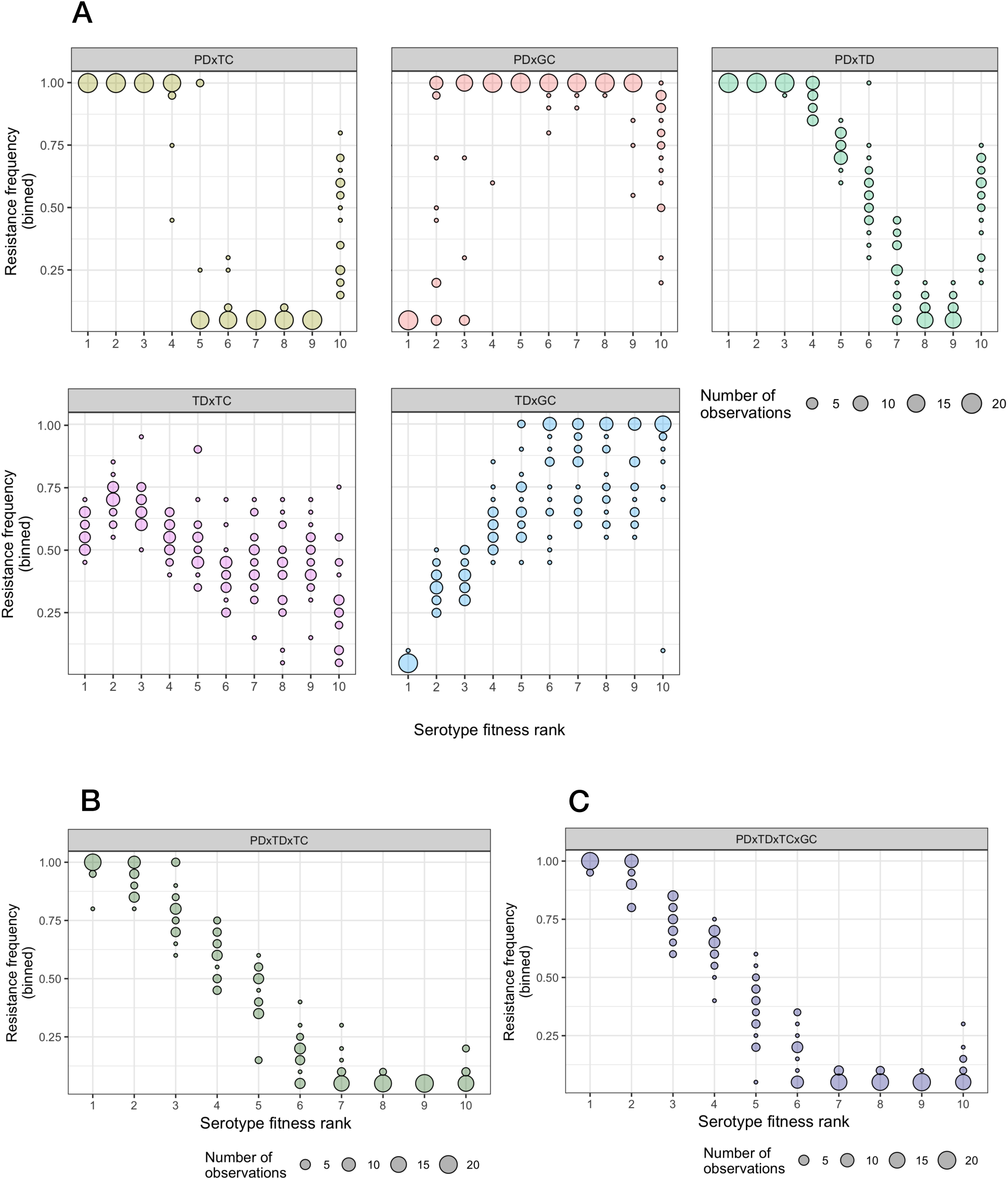
A range of serotype-specific resistance patterns is possible when combining mechanisms. Variation in resistance frequency of each hypothetical serotype from 20 model simulations of combinations of the four models. Resistance frequency (y axis) is binned, with the size of the circle indicating the number of repeated model simulations that fell within that bin. A) two-way combinations between the models, B) three-way, and C) four-way combinations of the models.s

In the treatment diversity model, sub-groups of the population with different treatment rates mix assortatively. To test of whether this differential mixing between subpopulations alone was enough to generate intermediate resistance frequencies in the combined models, we combined the treatment competition model and the treatment diversity model without any variation in treatment rate between groups (S. Fig 5). The serotype-specific resistant patterns under this combination were comparable to treatment competition model acting alone.

Finally, we tested whether a small within-host growth advantage for sensitive strains in addition to all other mechanisms was compatible with observed serotype-specific resistance patterns using a 4-way combination of all models. Within this model, there are two costs to resistance: a transmission cost and a within-host growth cost. We added 1/10 and 1/2 of the growth advantage sensitive strains needed to maintain the target resistance frequency when the growth competition was acting alone. Under this combination of models, both the empirically-observed positive association between serotype frequency and resistance frequency and within-serotype coexistence was maintained (Table 2, Fig 4C, S. Fig 6).

## 5 Discussion

Using empirical carriage data and individual-based multi-serotype models, we identified sets of mechanisms of sensitive-resistant strain competition that can capture pneumococcal resistance epidemiology. Of the mechanisms studied here, we show that both host population transmission structure and treatment heterogeneity were needed to explain coexistence of sensitive and resistant strains of the same serotypes; that serotype-specific carriage duration or within-host competition (or both) could explain differences in resistance frequency by serotype; and that differential within-host growth of sensitive and resistant strains may not be strong enough to play a role in population-level transmission dynamics. We highlight what further empirical data and analyses that are needed to further hone in on the strength of each mechanism that would best explain resistance dynamics.

Although within-host competition models have been shown to capture empirical resistance frequencies in single-serotype models, we show that serotype-level coexistence is lost or depleted when these models are extended to include multiple serotypes, so that these co-colonisation models alone could not replicate carriage data. We suggest this loss of serotype-level coexistence is due to host adaptive immunity, which blocks same-serotype co-colonisation that is required to generate the negative-frequency dependent selection which maintains the intermediate resistance frequency in the absence of multiple serotypes. Our argument is supported by our previous modelling work, which found more extensive intermediate resistance frequencies within serotypes when immunity is gained on clearance of a serotype rather than adaptive immunity being gained following colonisation (Davies et al., 2019). However, recent immunological evidence suggests that serotype-specific adaptive immunity is gained on acquisition, not clearance (Ramos-Sevillano et al., 2019). Our results highlight the potential importance of interactions between immune-mediated negative-frequency dependent selection (NFDS) and other NFDS processes that act on loci that impact strain fitness on pathogen diversity. In this case, two nested diversifying selection processes resulted in depletion rather than maintenance of within-serotype diversity.

Serotype-level co-existence seen in empirical data can be regained in the within-host competition co-colonisation model when it is combined with host population structure (via the treatment diversity model). The treatment diversity model permits coexistence of sensitive and resistant strains by creating niches for each serotype in the host population and thus overriding the effects of adaptive immunity. On its own, a population structure model was insufficient to explain why serotypes would have different resistance frequencies.

However, the combined model captured both the association between serotype carriage prevalence and resistance frequency and the within-serotype co-existence observed empirically. Integrating population structure into the pathogen diversity model worked in a similar way: by itself each mechanism explained one of the components of the empirical data, while combined they explained both. It is well established in ecological theory that niche structure (for example spatial structure) can promote coexistence of one trait (Chesson, 2000), but we are not aware of applications to explain nested diversity (such as sensitive and resistant strains coexisting within serotypes) in pathogens.

Our results show that faster transmission of some serotypes could explain the positive association between serotype prevalence and resistance frequency. This finding is consistent with previous studies (Davies et al., 2019). That serotypes or pneumococcal genomes with longer carriage duration have a higher resistance frequency is well established in theory and carriage studies (Lehtinen et al., 2017; Andrejko et al., 2022; Lees et al., 2017). However, to our knowledge, there has not been an analysis of loci associated with faster transmission and resistance frequency in pneumococcal genomes. PCV changes the transmission rates of serotypes, and it has been shown theoretically that this could impact resistance dynamics post-PCV (Davies et al., 2021). Empirically it is observed that resistance frequency increases in non-vaccine serotypes (Lo et al., 2019; Andrejko et al., 2021), which supports the theory that transmission could affect strength of selection for resistance. It is therefore crucial to show how influential transmission rates are on serotype-level resistance selection to accurately predict resistant carriage changes after vaccine introduction.

Our results suggest that a within-host growth resistance cost alone cannot capture patterns in pneumococcal resistance data, nor when combined only with carriage duration diversity or host population structure by treatment group. The growth competition model on its own and in 2-way combinations predicts a negative relationship between serotype prevalence and resistance frequency, which has not been empirically observed. However, sensitive strains can outcompete resistant strains due to growth rate-related fitness costs in vitro (Melnyk et al., 2015). How are these two observations compatible? Here we propose one solution by including a weak effect of differential growth alongside other strain competition mechanisms and a transmission resistance cost. When the growth differential is small, treatment usually occurs before within-host competition has resulted in loss of resistant strains, so that this process has little impact on the overall force of infection of sensitive and resistant strains, and is therefore not driving NFDS for sensitive strains. An alternative consideration that we have not addressed is that serotypes themselves differ in their within-host growth rate, as it has been shown that serotypes that are more competitive in vitro tend to be more resistant in carriage data (Andrejko et al., 2022). To what extent mathematical models of infectious disease need to account for both within-host and between-host scales to explain population-level trends is an ongoing question (Mideo et al., 2008; Childs et al., 2019).

Although we show that combining models is essential to capture all qualitative patterns in serotype-specific resistance data, we do not know how strongly each mechanism could be driving resistance patterns. It is likely that several different combinations of model strengths could explain observed data equally well, and that the parameters controlling the strength of each driver are unidentifiable. Larger datasets on community carriage that report serotype and antibiotic susceptibility would reduce uncertainty and permit more validation. In addition, data on co-colonisation would narrow down the set of likely combinations by adding data specific to the within-host model mechanisms, while controlled observations after an intervention may also provide insights (Davies et al., 2021).

## 6 Methods

### 6.1 Search strategy, study selection criteria, and data analysis

To identify studies reporting serotype-specific prevalence and penicillin susceptibility, we restricted our search to those included in a recent systematic review and meta-regression of carriage and invasive disease (Andrejko et al., 2021) that had already undergone comprehensive screening and quality assessment. We further restricted studies to those reporting serotype-specific prevalence and penicillin susceptibility in nasopharyngeal carriage isolates collected from children, prior to widespread introduction of pneumococcal conjugate vaccines (PCVs) in the population. We then excluded studies where participants had a mean age of less than 10 years old, and where children had been recruited because they were immunocompromised or had had a diagnosed respiratory condition.

For each study, we extracted the total number of samples taken, the total number of pneumococcal positive samples, the fraction of susceptible (i.e. sensitive) and non-susceptible isolates, the minimum inhibitory concentration (MIC) threshold used to indicate non-susceptibility and/or resistance in the study, country, region, information on age of cohort, and percentage of children vaccinated (as some children were vaccinated in the private market in the absence of national immunisation programmes).

Within each study, we then classified each pneumococcal positive isolate according to serotype or serogroup where serotype was not available, and penicillin susceptibility (with non-susceptible isolates broken down into intermediate or resistant isolates, where possible). From these data, for each serotype, we calculated 1) the serotype frequency (the fraction of isolates in the study belonging to that serotype), 2) the frequency rank (in decreasing order), and 3) the non-susceptibility frequencies (the fraction of isolates below the resistance cut off).

To understand the general association between non-susceptibility to penicillin and other variables, we fitted four binomial generalised linear mixed-effects models to the full dataset: 1) the null model, (intercept-only), 2) the serotype rank model (serotype frequency rank as a fixed effect), 3) the serotype class model (serotype identity as a random effect), and 4) the serotype class and rank model (serotype frequency rank as a fixed effect and serotype identify as a random effect). All models included study random effects to account for variation in susceptibility across settings, and an observation-level random effect, to deal with overdispersion. We used Akaike Information Criteria (AIC) to determine the best-fitting model.

### 6.2 Mathematical models

#### 6.2.1 Description

We developed a suite of four individual-based transmission models for each of the coexistence mechanisms previously shown to plausibly explain resistance frequency identified in Davies et al. (2021), and based on the individual-based pneumococcal transmission model described in Davies et al. (2019). A full description of the models is provided in the Supplementary Materials. In brief, we first constructed a base model for *L* serotypes, with one sensitive and one resistant strain for each subtype (*L* ∗ 2 = *M* strains in total). Each serotype is denoted *l* ∈ {1*, . . . , L*} and each strain is denoted *j* ∈ {1*, . . . , M* }. An individual, *i* ∈ {1*, . . . , N* }, has a continuous colonisation state with respect to the proportion of the colonisation niche taken up by each strain (*f_ij_*), a total colonisation status which is always 0 or 1 (*F_i_* = ^L^ *f_ij_*), and a serotype-specific immune state (*m_l_*) of 0 or 1. We characterised the following events: 1) transmission of strain *j* of serotype *l* and simultaneous gain of immunity to serotype *l*, which occurs in immunologically naive hosts at rate *κ^Fi^ λ_j_/N* , where *κ* is the relative rate of secondary colonisation to primary colonisation, and *λ_j_* is the strain-specific force of infection, based on a serotype-specific transmission rate *β_l_* and the sum of all colonisations of strain *j* in the population; 2) natural clearance, in which all strains are simultaneously lost, which occurs at rate *µ*, which is not serotype-specific in the base model; 3) immune waning, in which all immunity is lost, which occurs at rate *ϕ*; and 4) death or ageing of the individual, in which the colonisation status of individual *i* is reset, representing death or ageing out of the tracked population *br* and replacement with a fully susceptible newborn. As demonstrated in Davies et al. (2019), this base model is structurally neutral with respect to sensitive and resistant strains of each serotype (Lipsitch et al., 2009), so that all coexistence between sensitive and resistant is explained by the inputted mechanisms. Events are modelled as inhomogeneous Poisson processes. We only track colonisations in children (average age of 10 years old) to parsimoniously capture the majority of transmission events, which occur between children (Qian et al., 2022; Van Zandvoort et al., 2024), and to match carriage data.

Next, we incorporated four coexistence mechanisms into separate versions of this base model: treatment diversity (TD), pathogen diversity (PD), treatment competition (TC), and growth competition (GC) (Table 1, Supplementary Materials). We add antibiotic treatment events, which remove sensitive strains from hosts at rate *τ* for all models, although we do not expect clearing pneumococcal colonisation to be the target of antibiotic treatments. We also included a cost of resistance, as a reduction in transmission of the resistant strain for TD, PD, and GC models (*c*), relative to the susceptible strain, and as a benefit to growth of sensitive strain in co-colonisations in the GC model (*ω*). Co-colonisation is not possible in TD and PD models (*κ* = 0).

We also combined the models. When the treatment or growth competition models were combined with the pathogen diversity model, hosts may be colonised with more than one serotype and therefore subject to multiple serotype-specific clearance rates. Therefore, we adapted the model so that the clearance rate was a weighted average of the serotype-specific clearance rates of the serotypes with which they are currently colonised, ∑ *f_i_µ_l_*. We obtained 20 repeated simulations of each model individually, and for 2-, 3-, and 4-way combinations.

#### 6.2.2 Model parameterisation and calibration

First, we calibrated the transmission rates of each serotype in the base model without sensitive and resistant strain dynamics to the relative serotype-specific prevalence in the target dataset. We used a model emulator approach (Perumal and Zyl, 2025) that fitted a Gaussian Process model to the serotype prevalence ratios from model simulations obtained for different transmission rate ratios. We then used the emulator model to estimate the transmission rate ratios needed to obtain the serotype prevalence ratios observed in the data (see Supplementary Materials for full details).

Next, we fit each individual model to an overall resistance frequency by estimating the cost of resistance in each model. We used Bayesian Optimization (Pelikan, 2005) to find the resistance cost that minimised the distance between observed and model obtained resistance frequency (see Supplementary Materials for full details). For the PD model, it is possible to exactly estimate the resistance cost thresholds at which each serotype will be resistant, based on the treatment rate and the carriage duration of each serotype. We generated a resistance frequency for the mid-point between each threshold through model simulation, and selected the resistance cost that gave the most similar resistance frequency to the data.

Parameters that were not related to the strain competition mechanism (including average carriage duration and duration of immunity) were drawn from the literature and previous model fits (S.Table 2) and did not differ between models.

### 6.3 Software, data, and code availability

We conducted data and statistical analyses using R v4.3.2 (R Core Team, 2023) and the lme4 (Bates et al., 2015), DHARMa (Hartig, 2024) and sjPlot (LÃijdecke, 2024) packages. The individual-based models were implemented in Julia v1.11 (Bezanson et al., 2017), with the calibration and fitting using the GaussianProcesses (Fairbrother et al., 2022) and BayesianOptimization (Brea, 2021) packages.

## Supporting information

supplementary mats

## Data Availability

All data produced in the present study are available upon reasonable request to the authors

## 7 Acknowledgements

KEA, HCL, JH and EP-H were supported by the Wellcome Trust [219797/Z/19/Z]. KvZ was supported by the Bill and Melinda Gates Foundation [OPP1139859]. SF was supported by the Einstein Foundation Berlin as an Einstein BUA Strategic Professor [EPP-BUA-2022-697].

Kynan Delaney for helpful statistical discussions.

