## supplementary mats for "Multiple mechanisms of strain competition are needed to explain pneumococcal serotype-specific antibiotic resistance patterns"

### 1 Model description

We use a suite of individual-based, multi-serotype models of transmission of and immunity to *S. pneumoniae*. We track the fraction of colonisation taken up by sensitive and resistant strains within each host, with co-colonisation between any combination of strains possible. Hosts gain immunity on colonisation with a serotype which inhibits further colonisation by that serotype for the duration of immunity. The base model is structurally neutral with respect to sensitive and resistant strains of each serotype (see Davies et al, 2019). In the first section of the supplementary model description, we describe the base model. The second section describes the four coexistence mechanisms that were built into the base model. The third section describes parameterisation.

#### 1.1 Base model

##### 1.1.1 Strains and serotypes

Serotypes refer to the immunological groupings for pneumococcus, such as 6A, 23F etc. Each serotype has two strains, one that is sensitive and one that is resistant. The number of serotypes included in the model is $L$, and the total number of strains is $2L = M$. Serotypes are indicated with and $l \in \{1, \ldots, L\}$, and strains are indicated with $j \in \{1, \ldots, M\}$. The serotype that a strain belongs to is $ceiling(j/2)$.

##### 1.1.2 Timesteps

We use discrete time, progressing by $dt$ at each step. Timesteps varied by model (see S Table 2), and the unit used throughout is years.

##### 1.1.3 Event types and event rate calculation

Transmission, clearance, treatment, and waning of immunity events are modelled as inhomogeneous Poisson processes. At each time step, the total number of events of a given type in the whole population is generated as a draw from a Poisson distribution with rate equal to the event rate multiplied by the time step and population size, e.g. $\mu \cdot dt \cdot N$ for clearance events.

##### 1.1.4 Host colonisation and immune status

The colonisation of host $i$ by strain $j$ is denoted $f_{ij}$, with $f_i = (\ldots)$ giving the vector of colonisations for each type, and $F_i = \sum_{j=1}^{M} f_{ij}$ giving the sum of colonisations within the host.

The immune status of the host is denoted by $m_{il}$, with $m_i = (\ldots)$ giving the vector of the host immune status to each serotype. $m_{il} = 1$ if the host is immune to serotype $l$, and 0 otherwise.

##### 1.1.5 Transmission and immunity

The force of infection is calculated as $\lambda_j = \beta_l \sum_{i=1}^{N} f_{ij} dt$, where $\beta_l$ is the serotype-specific transmission rates. This gives the total expected number of exposure events (rather than successful colonisations) in a population per time step. In order to prevent stochastic loss of rare strains without facilitating coexistence, if $\lambda_j = 0$ it is set to a low value, $ymin = N/(10^5)$.

Hosts gain a colony of size $\iota$ of the transmitting strain following a successful transmission event. After any event that alters the colonisation value of some but not all strains (i.e., colonisation or treatment), there is a normalisation step which ensures that $F_i = 1$. Here, the new value of each colonisation is: $f_{ij} = \frac{f_{ij}}{(\sum_{j=1}^{M}(f_{ij}))}$. Therefore we can write the rate at which a host $i$ is newly colonised by strain $j$ as:

$$f_{ij} = 0 \xrightarrow{(1-m_{il})\kappa^{F_i}\lambda_j/N} f_{ij} = \frac{\iota}{(F_i + \iota)}$$

In the case of primary colonisation of an immunologically naïve host, this is equivalent to:

$$f_{ij} = 0 \xrightarrow{\lambda_j/N} f_{ij} = 1$$

On successful colonisation, immunologically naïve hosts gain immunity to the serotype $l$ of the colonising strain $j$, i.e.:

$$m_{il} = 0 \xrightarrow{\kappa^{F_i}\lambda_j/N} f_{ij} m_{il} = 1$$

All immunity wanes simultaneously at rate $\phi$:

$$m_{il} = 1 \xrightarrow{\phi} m_{il} = 0$$

##### 1.1.6 Clearance

All strains are cleared simultaneously in the host when there is a clearance event. Clearance occurs at rate $\mu$, i.e.

$$F_i = 1 \xrightarrow{\mu} F_i = 0$$

##### 1.1.7 Birth, death, and ageing

Hosts die or age out of the group of interest at rate b. After death, the host is immediately replaced by a newborn host with no colonisation, immune, or vaccine protection.

#### 1.2 Coexistence mechanisms

In all coexistence mechanisms below, we add a fitness cost to the resistant strain, and treatment events that remove sensitive strains from the host. Each mechanism uses a different model structure that permits coexistence of both strains. The treatment competition and growth competition models were first proposed in Davies et al, 2019. The treatment diversity model is drawn from Krieger et al 2020 and Davies et al, 2021, and the pathogen diversity model was first described as the D-types model in Lehtinen et al, 2017.

##### 1.2.1 Treatment competition

In the treatment competition model, the cost of resistance is specified as a reduction in the rate of acquisition of the strain, i.e. the force of infection of resistant strains is $\lambda_R = (1-c)\lambda_j$.

All sensitive strains are cleared simultaneously when there is a treatment event. Treatment occurs at rate $\tau$. Therefore, during a treatment event sensitive strains, which are oddly numbered, $s \in (1, 3, ..., M)$ are set to 0:

$$f_{is} > 0 \xrightarrow{\tau} f_i s = 0$$

After treatment, the remaining colonisations are normalised. This results in an increase in the colonisation values of any resistant strains colonising the host, providing a fitness advantage to resistant strains co-colonising with sensitive strains in the presence of treatment.

##### 1.2.2 Growth competition

In the growth competition model, the cost of resistance is specified as a reduced fitness when competing with sensitive strains within the host. The colonisation status of each type within a host is updated once per timestep. In hosts with mixed-susceptibility co-colonisations, sensitive strains increase their colonisation level and resistant strains decrease. Resistant strains are cleared during within-host growth if the fraction of colonisation falls below $f_{min}$. For sensitive strains $s \in (1, 3, \ldots, M)$, and resistant strains are $r \in 2, 4 \ldots, M$:

$$f_{is} = \frac{f_{is}\omega^{dt}}{\sum\limits_{\substack{s=1 \\ s \text{ odd}}}^{M} f_{is}\omega^{dt} + \sum\limits_{\substack{r=2 \\ r \text{ even}}}^{M} v(f_{ir})}$$

Where:

$$v(f_{ir}) = \begin{cases} f_{ir} & \text{if} f_{ir} > f_{min} \\ 0 & \text{if} f_{ir} < f_{min} \end{cases}$$

Treatment events work in the same way in the growth competition model as they do in the treatment competition model, i.e. removing all sensitive strains. The growth competition uses a smaller time step than other models, $dt = 0.5/365$.

##### 1.2.3 Treatment diversity

Under this coexistence mechanism, we split the host population into treatment groups, which each have a different treatment rate, and slower transmission between groups than within groups, and a transmission cost of resistance. We set $k = 0$ so that co-colonisation is not possible and hosts only carry one strain at a time, with no competition occurring between serotypes or strains within a host. Treatment groups are denoted $q \in 1, \ldots, T$, with $T$ treatment groups in total, with $N_T = \frac{N}{T}$ individuals per group. The colonisation status of strain $j$ in individual $i$ in group $q$ is $f_{ijq}$. There is greater transmission within treatment groups than between treatment groups, controlled by parameter $g$. The force of infection of strain $j$ in treatment group $q$ is:

$$\lambda_{jq} = \beta_j \left( g \frac{\sum_{i=1}^{N_T} f_{ijq}}{N_T} + (1-g) \sum_{\substack{u=1 \\ u \neq q}}^{T} \frac{\sum_{i=1}^{N_T} f_{iju}}{N_T} \right)$$

We assume that there is a Gamma distribution of treatment rates across these groups in the population. Treatment rates are drawn from a Gamma distribution with shape $\gamma$ and rate $\theta = \tau/\gamma$, where $\tau$ is the mean treatment rate in the population. We discretise the continuous gamma distribution by splitting it into $Q$ equal sized ordered treatment groups and finding the median treatment rate per group, by applying:

$$\tau_q = Quantile_{\Gamma(\gamma,\theta)}(\frac{q}{Q} - \frac{0.5}{Q})$$

The variation in treatment rates and slow mixing between groups results in parameter space in which both sensitive and resistant strains are maintained as population levels, with different frequencies of each strain in each treatment group.

###### 1.2.4 Pathogen diversity

In this model, there is a different clearance rate per serotype, i.e. for serotype $l$ the clearance rate is $\mu_l$, and a transmission cost for resistance, and no co-colonisation ($k = 0$). Serotypes with longer carriage durations are more exposed to more treatment events thereby experiencing stronger selection for resistance, so that serotypes with $\mu_l < \frac{\tau(1-c)}{c}$ are resistant (Lehtinen et al, 2017).

#### 1.3 Model sampling algorithm

To simulate from the model efficiently, we used the algorithm described in Davies et al, 2019. Once per timestep we updated event rates, estimating the total possible number of events of each type during the next time step. We drew actual numbers of events from inhomogeneous Poisson processes. Next, we randomised the order of these events, and randomly assigned a host to each event. Finally, we evaluated every event, first checking whether the event would occur (depending on host colonisation and immune status), and then applying it.

#### 1.4 Parameterisation

Where possible, we used estimates from epidemiological data for parameterisation. We also use estimates from previous modelling studies (S Table 1, S Table 2, S Table 3). We also use calibration for transmission rates ($\beta_j$) and resistance cost ($c$ or $\omega$).

##### 1.4.1 Calibration approach

We calibrate the model in two steps with the aim of achieving a realistic distribution of relative serotype prevalence and overall resistance frequency. We fit to serotype and resistance data in a large pneumococcal carriage survey (Espinosa-de Los Monteros et al, 2007).

##### 1.4.2 Transmission rates

To find transmission rates for each serotype, we simulated prevalence rate ratios for different transmission rate ratios from the base model with treatment, and fit a Gaussian Process model as an emulator of this relationship. Then, we extracted relative prevalence ratios for each of the 10 most prevalent serotypes in Espinosa-de Los Monteros et al, 2007, and used the to estimate the transmission rate ratios required to achieve these observations.

Setting the maximum transmission rate at 200, we simulated prevalence rate ratios for the least compared to the most prevalent serotypes in a model with five serotypes. This data was then used to fit the Gaussian Process model (S. Fig. 1A, S Table 3). A sensible fit was obtained between target relative serotype prevalence and the model output (S. Fig

1B).

##### 1.4.3 Cost of resistance

Next, we calibrated the cost of resistance to achieve a 66.7% overall resistance frequency. We used the frequency of nonsusceptibility to penicillin in isolates of *S. pneumoniae* from Espinosa-de Los Monteros et al, 2007. In the TD, TC, and GC models we used Bayesian Optimization (Pelikan, Goldberg and Cantu-Paz, 2005. Package: BayesianOptimization.jl v0.2 Brea, 2021) to minimise the difference between simulated and targetted overall resistance frequency. For the TD and TC models, we sampled 100 points from parameter space, and for the GC models, we sampled 50. We ran the model twice for each sampled parameter. From this we obtained a rough best estimate of the resistance cost needed to obtain the target resistance frequency (S Fig 2)

### 2 Supplementary results

#### 2.1 Data extraction

Of the 558 studies screened, we found 12 studies that met all inclusion criteria (3), covering 9,977 children from 12 countries and 15 collection periods between 1995 to 2019. A total of 9,977 children were sampled across all studies, of which 3,541 had a positive nasopharyngeal swab for *S. pneumoniae*. In total, 3,600 *S. pneumoniae* isolates were detected, as some studies reported co-colonisation with multiple serotypes (in 4/15 collection periods), although they did not disaggregate resistance susceptibility results by co-colonisation status.

In total, 50 different named serotypes were identified across the studies (10.6% of *S. pneumoniae* isolates were classified as non-typeable, grouped into low-frequency categories, or had serogroup-only information). Excluding unnamed isolates, each serotype was observed on average in 5 studies. Serotypes 19F, 23F, 6A, and 6B were observed in all studies, while thirteen serotypes were observed in one study each. The average number of isolates across all studies identified for a particular serotype was 64. Serotype 19F was the most prevalent serotype (597 total isolates) whereas 66% of serotypes (33 serotypes) were observed in less than 20 total isolates, and 13 serotypes (26%) were only observed in five or fewer isolates. On average, 16 different serotypes were observed per study (ranging from 7 in Romania to 33 in Mexico). Four serotypes took up the top-ranking spot across all included studies (19F, 23F, 6A, and 6B)).

Among serotypes reported in 5 or more studies, the serotype with the highest average resistance frequency was 6B, with a resistance frequency of 52.1% on average (variance 12.6). 16 serotypes were never recorded as being resistant, including 18F which was detected in 4 studies in a total of 15 isolates.

#### 2.2 Additional model results: Resistant strain invasion fitness

Although intermediate resistance frequencies are possible for each model across the whole pneumococcal population (S Fig 4), in the treatment and growth competition models there is a lack of coexistence of sensitive and resistant types within serotypes (Main text, Fig 3A, Table 2). We hypothesised that this loss of serotype-level coexistence is due to serotype-specific immunity leading to either the sensitive or the resistant strain of one serotype suppressing the other. To demonstrate and test this idea, we used a simplified two-serotype version of the within-host models, investigating what proportion of resistant colonisations that were taken up by each resistant strain in the two-serotype model, for different values serotype fitness (i.e., transmission rate) (S Fig 7A). In these models, we denote two serotypes A and B, and a sensitive and resistant version of each type (As, Ar, Bs, and Br). In these experiments, either one resistant type or the other tends takes up all the resistance niche, depending on which serotype is fitter. In the TC model, resistant strain fitness is higher when serotype fitness is higher. In the GC model, on the other hand, coexistence was possible when both serotypes had low transmission fitness or when one type was much less fit than the other.

We propose as an explanation for this behaviour that if one of the sensitive strains gains an advantage (e.g., As), either stochastically or through higher fitness, this leads to a double benefit to the resistant type of the opposite serotype (e.g., Br). Negative frequency-dependence gives increases transmission of the Br twice: being the rarer serotype, it encounters less immune blocking, and being a resistant type, it benefits from co-colonisations with the sensitive types (As). At the same time, As suppresses transmission of Ar through immune blocking, and Br suppresses transmission of Bs through immune blocking. This leads to a double suppression of the transmission of Ar: through immune blocking depleting immunologically naive hosts, and through reduced transmission of Bs, depleted opportunities for co-colonisation. In the GC model, on the other hand, if transmission of one or both serotypes is low enough that co-colonisation is very rare, the impact of immune suppression on the dynamics of resistant strains is weaker.

We further investigated this theory by introducing the Br strain as an invader, after As, Ar, and Bs were simultaneously introduced and reached equilibrium levels in the population (S Fig 7B). Under these conditions, the region in which Br can be the dominant resistant strain is reduced under both models, although to a much greater extent in the TC model. Even if Br has a fitness twice as high as Ar (through transmission rate), it cannot invade. This lends support to our theory as the extensive immunity generated by Bs and Ar in the population would simultaneously block transmission of Br and of its main resource for co-colonisation, As.

### 3 Supplemental tables and figures

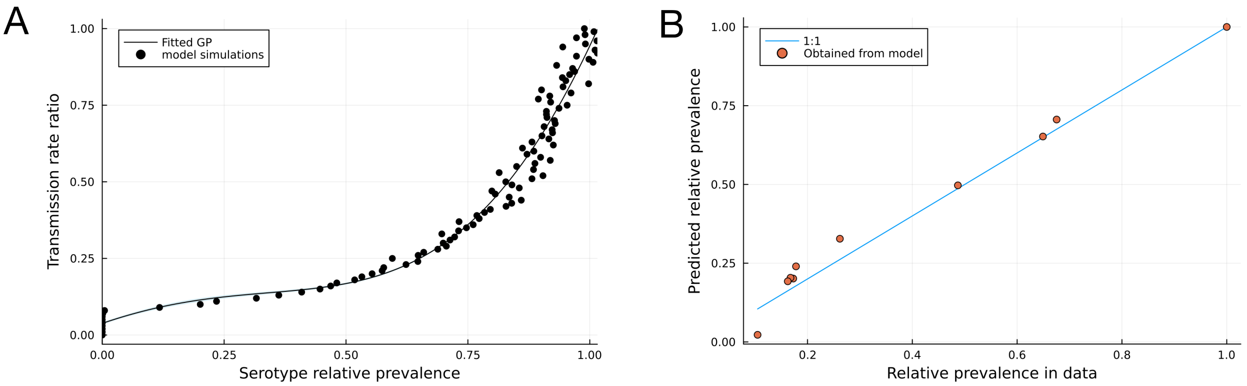


Figure 1: Calibrating transmission rate ratios. A) Relative prevalence rates against transmission rate ratios. Points are the model output, and the line and ribbon is the fitted Gaussian Process model. B) Target vs obtained relative serotype prevalences for the ten most prevalent serotypes.

Table 1: Parameter definitions and sources

| Parameter | Definition | Source |
|---|---|---|
| $\beta_{max}$ | Maximum serotype-specific transmission rate per capita per year | Calibrated to Espinosa-de Los Monteros et al, 2007 |
| $\beta_l$ | Serotype-specific transmission rate | Calibrated to Espinosa-de Los Monteros et al, 2007 |
| $\mu_l$ | Seroptype-specific rate of clearance of all colonisations per capita per year | Hill et al, 2008 |
| $\kappa$ | Reduction in rate of acquisition of colonisations in an already colonised host | Davies et al, 2021 |
| $\iota$ | Size of starting colonisation | Davies et al, 2021 |
| $f_{min}$ | Colonisation size below which strains are cleared %during within-host growth | Davies et al, 2021 |
| N | Population size | Cobey and Lipsitch, 2012 |
| L | Number of serotypes | - |
| br | Rate of death, birth, and ageing-out per capita per year | Set to make average host age 10yo |
| $\phi$ | Rate of waning of all immunity per capita per year | Flasche et al, 2013, Ravos-Sevillano, Ercoli and Brown, 2019 |
| $\tau$ | Average rate of colonisation-clearing treatments per capita per year | - |
| c | Reduction in transmission of resistant strains compared to sensitive strains | Calibrated to Espinosa-de Los Monteros et al, 2007 |
| $\omega$ | Relative growth rate of sensitive strains compared to resistant strains in mixed-susceptibility co-colonisations | Calibrated to Espinosa-de Los Monteros et al, 2007 |
| g | Relative amount of transmission occurring within vs. between treatment groups | Davies et al, 2021 |
| $\gamma$ | Shape parameter of gamma distribution of treatment rates | Davies et al, 2021 |
| T | Number of treatment groups | Davies et al, 2021 |

| Parameter | Base model | TD | PD | TC | GC | TDxPD | TDxTC | TDxGC | PDxTC | PDxGC | TDxPDx TC | TDxPDx TCxGC |
|---|---|---|---|---|---|---|---|---|---|---|---|---|
| $\beta_{max}$ | 200 | 200 | 200 | 200 | 200 | 200 | 200 | 200 | 200 | 200 | 200 | 200 |
| $\beta_l$ | see table 3 | see table 3 | see table 3 | see table 3 | see table 3 | see table 3 | see table 3 | see table 3 | see table 3 | see table 3 | see table 3 | see table 3 |
| $\mu_l$ | All 9.6 | All 9.6 | Linearly spaced between 7.2 and 12 | All 9.6 | All 9.6 | Linearly spaced between 7.2 and 12 | All 9.6 | All 9.6 | Linearly spaced between 7.2 and 12 | Linearly spaced between 7.2 and 12 | Linearly spaced between 7.2 and 12 | Linearly spaced between 7.2 and 12 |
| $\kappa$ | 0.95 | 0 | 0 | 0.95 | 0.95 | 0 | 0.95 | 0.95 | 0.95 | 0.95 | 0.95 | 0.95 |
| $\iota$ | 0.001 | 1 | 1 | 0.001 | 0.001 | 1 | 0.001 | 0.001 | 0.001 | 0.001 | 0.001 | 0.001 |
| $f_{min}$ | 3.00E-05 | 0 | 0 | 0 | 3.00E-05 | 0 | 0 | 3.00E-05 | 0 | 3.00E-05 | 0 | 3.00E-05 |
| N | 50,000 | 100,000 | 50,000 | 50,000 | 50,000 | 50,000 | 50,000 | 50,000 | 50,000 | 50,000 | 50,000 | 50,000 |
| L | 10 | 10 | 10 | 10 | 10 | 10 | 10 | 10 | 10 | 10 | 10 | 10 |
| br | 0.1 | 0.1 | 0.1 | 0.1 | 0.1 | 0.1 | 0.1 | 0.1 | 0.1 | 0.1 | 0.1 | 0.1 |
| $\phi$ | 1 | 1 | 1 | 1 | 1 | 1 | 1 | 1 | 1 | 1 | 1 | 1 |
| $\tau$ | 0 | 1 | 1 | 1 | 1 | 1 | 1 | 1 | 1 | 1 | 1 | 1 |
| c | 0 | 0.0844138 | 0.0984 | 0.102482 | 0 | 0.0844138 | 0.102482 | 0 | 0.102482 | 0 | 0.102482 | 0.102482 |
| $\omega$ | 1 | 1 | 1 | 1 | 1.06904 | 1 | 1 | 1.06904 | 1 | 1.06904 | 1 | 1.00690421 |
| g | 1 | 0.98 | 1 | 1 | 1 | 0.98 | 0.98 | 0.98 | 1 | 1 | 0.98 | 0.98 |
| $\gamma$ | NA | 3.5 | NA | NA | NA | 3.5 | 3.5 | 3.5 | NA | NA | 3.5 | 3.5 |
| T | 1 | 10 | 1 | 1 | 1 | 10 | 10 | 10 | 1 | 1 | 10 | 10 |

Table 2: Parameter values for each model

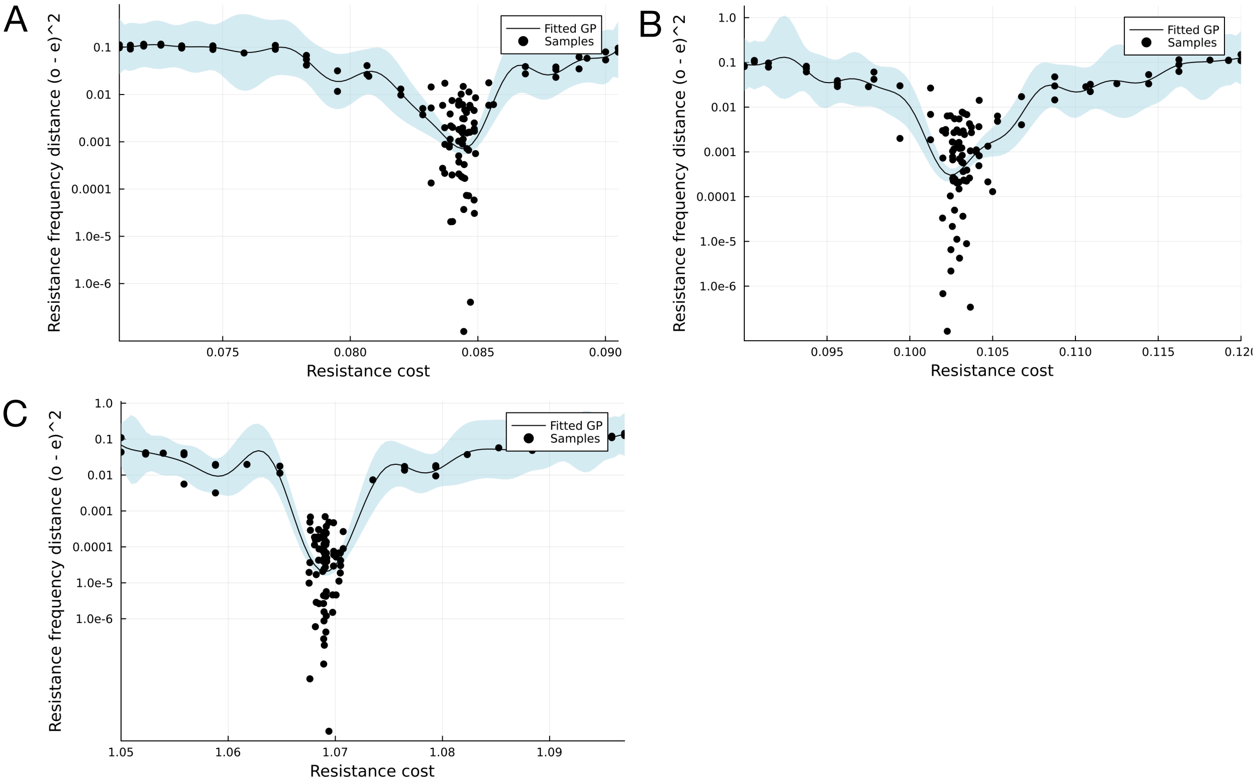


Figure 2: Calibrating resistance costs. Sampled parameter space in points and fitted Gaussian Processes model in line and ribbon, for the treatment diversity (A), treatment competition (B), and growth competition (C) models.

Table 3: Serotype-specific relative prevalence to most common serotype, drawn from Espinosa-de Los Monteros et al, 2007, and transmission rates obtained from calibration

| **Serotype** | **Relative ratio** | **Transmission rate** |
|---:|---:|---:|
| 1 | 1.00 | 200.00 |
| 2 | 0.68 | 54.17 |
| 3 | 0.65 | 49.17 |
| 4 | 0.49 | 31.39 |
| 5 | 0.26 | 25.78 |
| 6 | 0.18 | 23.16 |
| 7 | 0.17 | 22.93 |
| 8 | 0.17 | 22.68 |
| 9 | 0.16 | 22.42 |
| 10 | 0.10 | 18.77 |

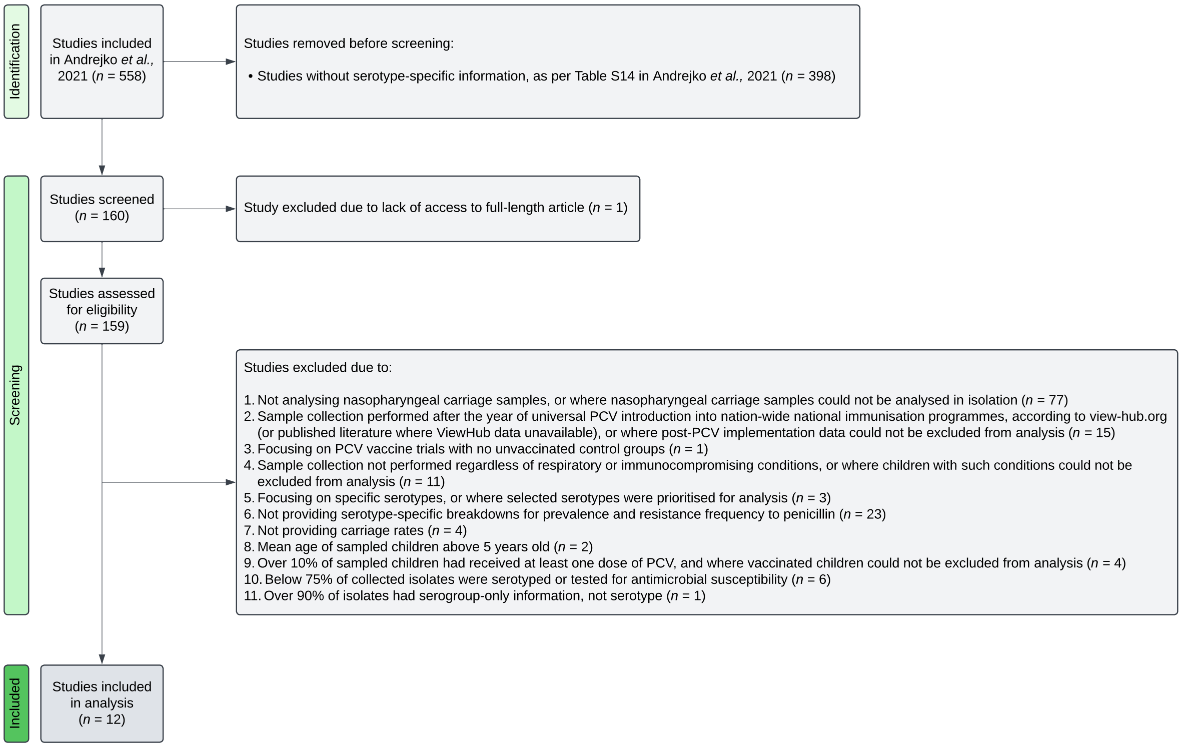


Figure 3: Flow diagram outlining the study selection process, with stepwise exclusion criteria.

Table 4: Random-effects logistic regression models for penicillin nonsusceptibility frequency.

| | **Base model** | | | **Rank model** | | | **Serotype model** | | | **Rank + serotype model** | | |
|---|---|---|---|---|---|---|---|---|---|---|---|---|
| *Predictors* | *Odds Ratios* | *CI* | *p* | *Odds Ratios* | *CI* | *p* | *Odds Ratios* | *CI* | *p* | *Odds Ratios* | *CI* | *p* |
| (Intercept) | 0.18 | 0.05 – 0.71 | **0.014** | 0.01 | 0.00 – 0.11 | **<0.001** | 0.13 | 0.03 – 0.54 | **0.005** | 0.03 | 0.00 – 0.22 | **0.001** |
| rank | | | | 1.10 | 1.04 – 1.16 | **<0.001** | | | | 1.07 | 1.01 – 1.14 | **0.031** |
| **Random Effects** | | | | | | | | | | | | |
| $\sigma^2$ | 3.72 | | | 3.68 | | | 2.90 | | | 3.09 | | |
| $\tau_{00}$ | 3.45 $_{\text{observation}}$ | | | 3.40 $_{\text{observation}}$ | | | 2.62 $_{\text{observation}}$ | | | 2.82 $_{\text{observation}}$ | | |
| | 6.85 $_{\text{study}}$ | | | 7.28 $_{\text{study}}$ | | | 0.88 $_{\text{serotype}}$ | | | 0.55 $_{\text{serotype}}$ | | |
| | | | | | | | 6.86 $_{\text{study}}$ | | | 7.10 $_{\text{study}}$ | | |
| ICC | 0.65 | | | 0.66 | | | 0.73 | | | 0.71 | | |
| N | 293 $_{\text{observation}}$ | | | 293 $_{\text{observation}}$ | | | 293 $_{\text{observation}}$ | | | 293 $_{\text{observation}}$ | | |
| | 15 $_{\text{study}}$ | | | 15 $_{\text{study}}$ | | | 15 $_{\text{study}}$ | | | 15 $_{\text{study}}$ | | |
| | | | | | | | 52 $_{\text{serotype}}$ | | | 52 $_{\text{serotype}}$ | | |
| Observations | 293 | | | 293 | | | 293 | | | 293 | | |
| Marginal $R^2$ / Conditional $R^2$ | 0.000 / 0.648 | | | 0.046 / 0.680 | | | 0.000 / 0.728 | | | 0.023 / 0.719 | | |
| AIC | 784.315 | | | 772.676 | | | 771.134 | | | 768.532 | | |

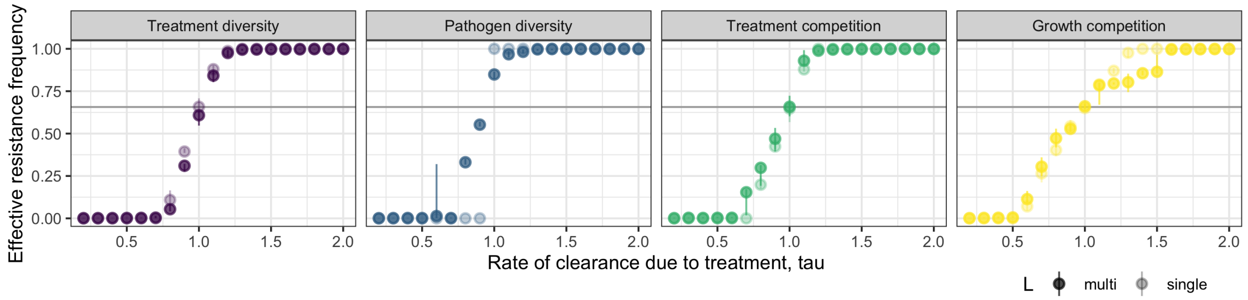


Figure 4: Resistance frequency for different values of treatment rate, showing the size of the window of coexistence, where intermediate resistance frequencies are found. Circle indicates the mean of 10 repeats, with error bars indicating 95% quantiles. Solid filled circles are for the full model with 10 serotypes, light filled circles are for the same model with one serotype.

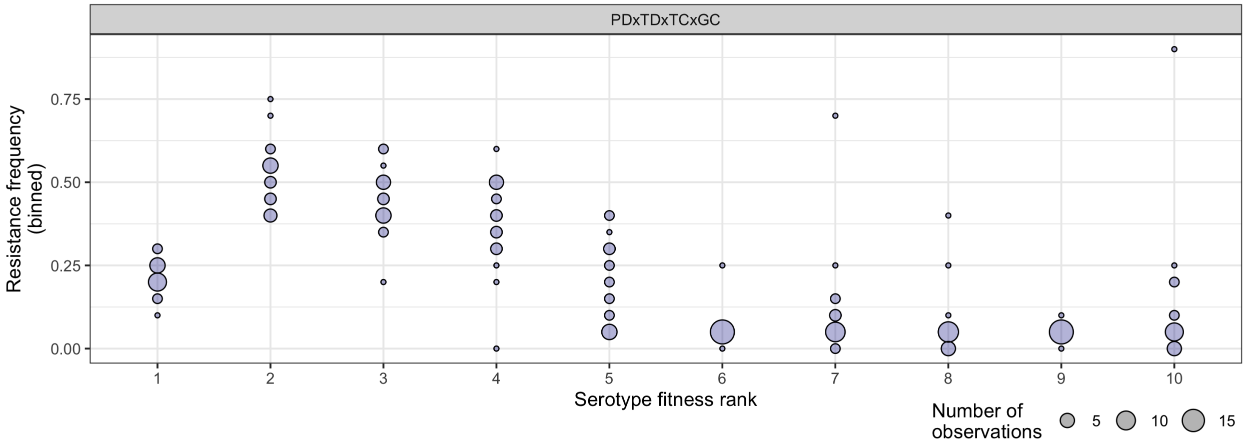


Figure 5: Variation in resistance frequency of each hypothetical serotype from 20 model simulations of all four models combined, with 1/2 of the growth advantage in sensitive strains required to maintain observed resistance frequency when the growth competition model acts alone. Resistance frequency (y axis) is binned, with the size of the circle indicating the number of repeated model simulations that fell within that bin.

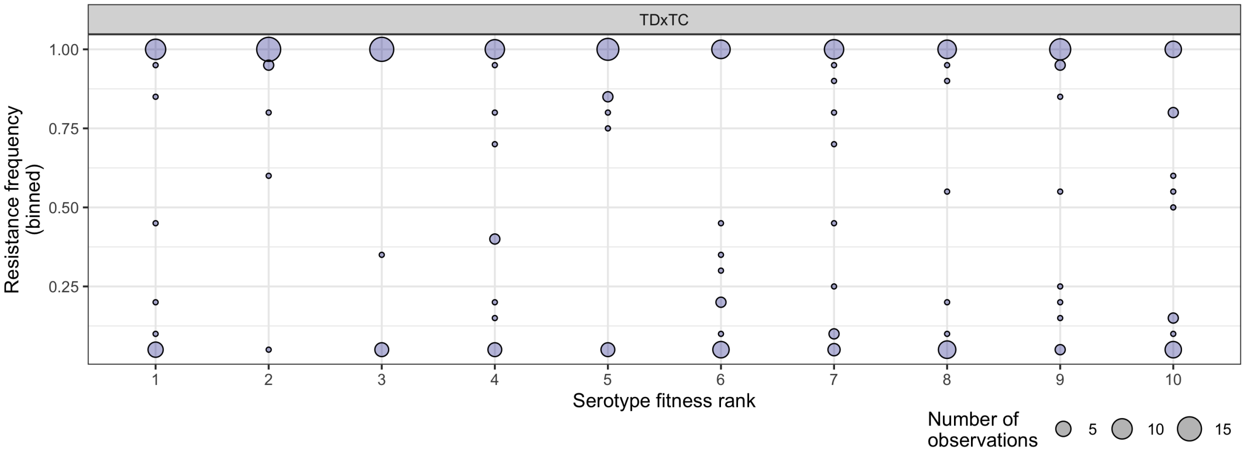


Figure 6: Variation in resistance frequency of each hypothetical serotype from 20 model simulations of the treatment competition model with the metapopulation structure of the treatment diversity model. Resistance frequency (y axis) is binned, with the size of the circle indicating the number of repeated model simulations that fell within that bin.

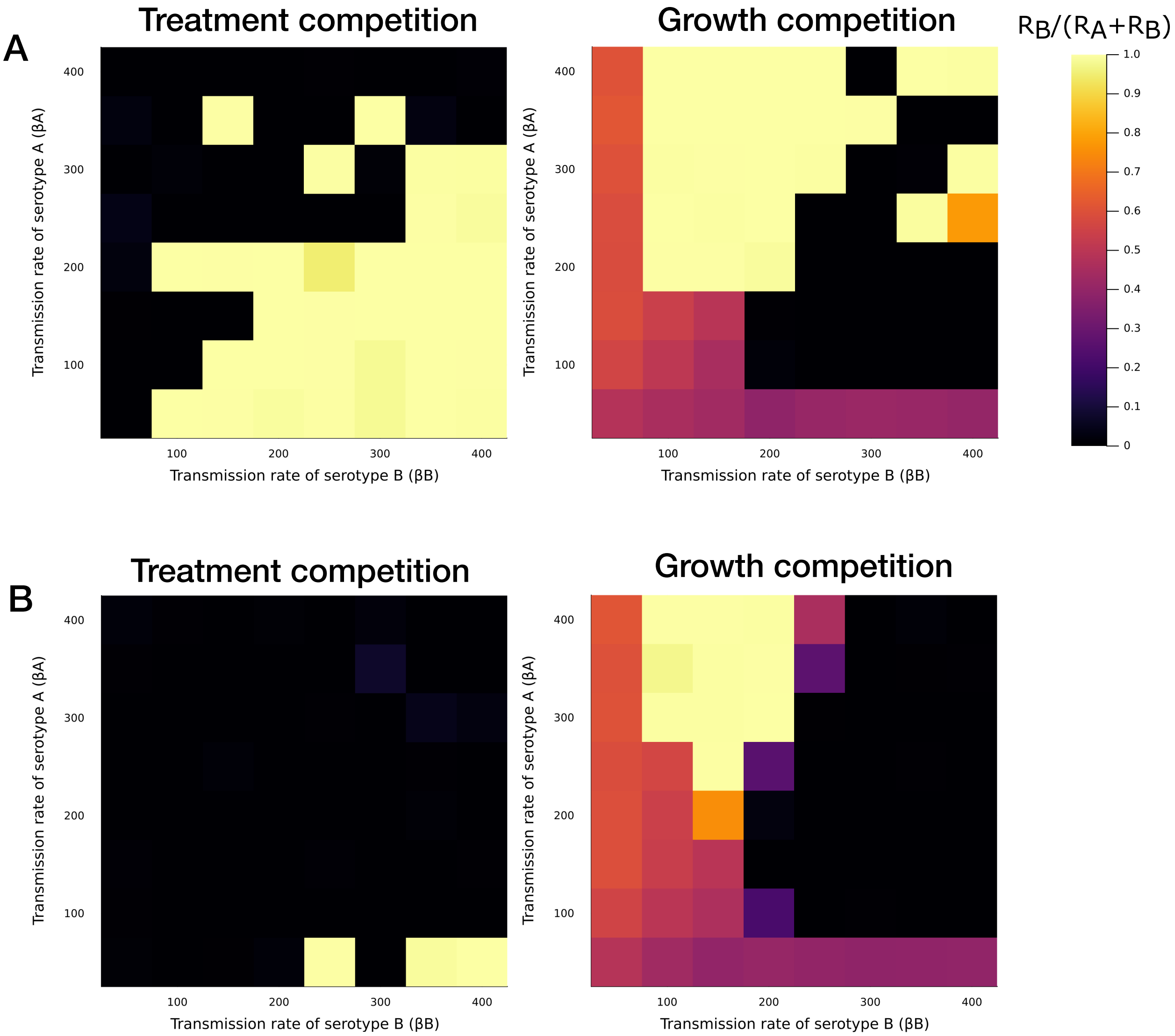


Figure 7: **Impact of immune competition on resistant strain dynamics in within-host competition models.** Proportion of resistant colonisations taken up by the strain Br in the two-serotype version of the TC and the GC model. In the top row, all four strains are introduced at the same time. In the bottom row, Br is introduced as an invader.
